# Autoimmunity and Arthritis in Youth with Autism and Suspected Post-Infectious Deteriorations

**DOI:** 10.64898/2026.03.19.26348838

**Authors:** Meiqian Ma, Noelle Schlenk, Jesse Sandberg, Zadie Schaffer, Kate Miles, Cindy Manko, Bahare Farhadian, Kayla Azad, Christy Capestany, Abhinay Aeruva, Yuhuan Xie, Paula Tran, Melissa Silverman, Kevin W Hoffman, Margo Thienemann, Jennifer Frankovich

## Abstract

The causes of severe neuropsychiatric deteriorations among patients with previously stable autism spectrum disorder (ASD) are poorly understood and present substantial challenges for care. We aimed to characterize the prevalence of autoimmune and inflammatory conditions and markers, as well as musculoskeletal findings, among youth with ASD experiencing a suspected post-infectious neuropsychiatric deterioration. The Stanford Immune Behavioral Health (IBH) Clinic is a specialty program for youth with neuropsychiatric deteriorations that are suspected to be post-infectious (non-psychosocial). We report findings for 43 consecutive patients with ASD (70% male [30 of 43]) evaluated in the IBH Clinic. The average (SD) age at clinical presentation was 12.0 (4.0) years. Juvenile arthritis was diagnosed in 15 patients (35%), predominantly enthesitis-related arthritis (ERA) and psoriatic arthritis (PsA). Seven patients had ultrasonographic evidence of joint effusions and/or synovitis without meeting juvenile idiopathic arthritis (JIA) criteria. Autoimmune conditions other than arthritis were observed in 9 patients (21%). The mean (SD) age at arthritis and other autoimmune condition diagnoses were 16.2 (5.5) and 12.7 (4.9) years, respectively. We observe markers of immune activation during neuropsychiatric deteriorations in over half of patients (60% [26 of 43]), including markers of autoimmunity (33% [12 of 36]), complement activation (41% [13 of 32]), immune dysregulation/inflammation (11% [4 of 37]), and vasculopathy (30% [13 of 43]). One-third (37% [16 of 43]) demonstrated two or more markers. These data underscore the importance of targeted immune evaluation—including musculoskeletal imaging and inflammatory marker screening—in ASD patients who have had a suspected post-infectious behavioral regression.

**Lay Summary:** In this cohort study of 43 patients with autism spectrum disorder (ASD) and suspected post-infectious deteriorations, more than half had laboratory markers of immune activation (using a limited panel), one-third had joint inflammation (confirmed by ultrasound), and additional autoimmune conditions were observed in 21%. From this, we conclude that patients with ASD who experience a suspected post-infectious neuropsychiatric deterioration may have underlying inflammation which may contribute to neuropsychiatric and behavioral regressions, highlighting the importance of immunologic and rheumatologic evaluation in clinical assessment.

## 1 | INTRODUCTION

Children with autism spectrum disorder (ASD) experience neuropsychiatric deteriorations – periods of symptom escalation and functional decline – at substantially higher rates than their neurotypical peers [1,2]. These deteriorations can be severe and often require acute psychiatric care, with inpatient admissions occurring at several-fold higher rates than youth without ASD [3]. Such deteriorations carry a heavy burden for families and the healthcare system [4]. Identifying the drivers of these deteriorations is uniquely challenging in ASD, as communication barriers and altered sensory awareness can obscure the reporting of symptoms [5–7]. Objective clinical evaluations are therefore critical.

Immune and inflammatory dysregulation is increasingly linked to ASD and represents an important area of investigation for both ASD etiology and treatment development [8–10]. These immune disturbances can present with physical manifestations such as arthritis and other rheumatologic illness, which may in turn contribute to pain, fatigue, and functional decline.

We present a retrospective analysis of patients with ASD evaluated in the Stanford Immune Behavioral Health (IBH) Clinic, a multidisciplinary program integrating rheumatology, immunology, and psychiatry. Previously, we have demonstrated that arthritis and autoimmune conditions occur with increased frequency among children with pediatric acute-onset neuropsychiatric syndrome (PANS) [11]. In the present study, we likewise evaluate rheumatologic illness (including arthritis) and signs of immune activation in youth with ASD who present after a neuropsychiatric deterioration. We postulate that immune activation and arthritis represent important, under-recognized contributors to pain and functional impairment in this population, with implications for both evaluation and treatment. Additionally, the impact of systemic inflammation on the blood-brain barrier and neuronal signaling may contribute to behavioral and cognitive decline.

## 2 | METHODS

### 2.1 | Study Design and Participants

Patients are referred to the IBH Clinic in cases where the referring provider is suspicious for a post-infectious or immunologically mediated neuropsychiatric deterioration. We reviewed charts of 499 consecutive patients evaluated between September 15, 2012, and January 31, 2024. Of these, 43 had ASD and are included in this study. ASD diagnoses were verified by a mental health clinician (CM) through retrospective chart review, confirming that the diagnosing provider (psychiatrist, psychologist, or developmental pediatrician) had appropriate expertise in ASD. Study data were collected and stored using REDCap (Appendix A.1) [12,13]. Sex and race/ethnicity were self-reported and included to assess generalizability. Race/ethnicity categories used were those from the Office of Management and Budget (2024 revision) [14].

### 2.2 | Characterization of Neuropsychiatric Deteriorations

We characterize neuropsychiatric deteriorations as escalations of psychiatric symptoms and heightened functional impairment relative to a patient’s baseline function [15]. Characteristics of each deterioration were obtained through review of the medical records and patient questionnaires (Table A1). Unclear dates were estimated using specific documentation protocol (Appendix A.2).

### 2.3 | Arthritis Classification

Rheumatologists’ records were used to assess joint pain, stiffness, warmth, swelling, redness, entheseal/joint tenderness, nail pitting, and signs/symptoms of inflammatory back pain (IBP) (by Calin criteria) [16]. Pediatric rheumatologists (MM, JF) classified patients with arthritis if they had (1) pain and joint effusion lasting greater than six weeks; or (2) pain and two or more of the following lasting greater than six weeks: limited or painful range of motion, tenderness, or warmth; or (3) joint tenderness and ultrasonographic confirmation of arthritis. The first two categories meet American College of Rheumatology (ACR) juvenile idiopathic arthritis (JIA) criteria. Patients were further classified by the International League of Associations for Rheumatology (ILAR) and/or Assessment of SpondyloArthritis International Society (ASAS) criteria (Appendix A.3) [17,18].

### 2.4 | Musculoskeletal Ultrasonography

Specialized musculoskeletal ultrasound of hands, wrists, feet, and ankles were obtained if (1) pain and stiffness persisted despite non-steroidal anti-inflammatories; (2) suspicion for arthritis was high based on history but physical examination findings were not definitive; and/or (3) the history or physical examination was incomplete due to psychiatric symptoms. We report specifications of ultrasound technology (Appendix A.4).

### 2.5 | Autoimmune and Other Immunologic Conditions

Diagnoses of celiac disease, psoriasis, thyroiditis, inflammatory bowel disease (IBD) (including Crohn’s disease and ulcerative colitis), and systemic lupus erythematosus (SLE) were assigned based on sub-specialist evaluation (Appendix A.5). Separately, we evaluated “other immunologic conditions” including asthma, seasonal or environmental allergies, food allergies, eosinophilic esophagitis (EoE), and primary immunodeficiency (PID) (Appendix A.5). We report the prevalence of food sensitivities, but we do not count this as an immunologic condition because it is often explained by sensory dysregulation.

### 2.6 | Laboratory and Physical Signs of Immune Activation and Anemia

We evaluated immune activation signs based on a predetermined set of laboratory and physical examination findings indicative of (1) autoimmunity (anti-histone antibody [high or normal], anti-thyroglobulin antibody [high or normal], anti-thyroid peroxidase [high or normal]); (2) complement activation (C1q binding assay [high or normal], complement C3 [low or normal], and complement C4 [low or normal]); (3) immune dysregulation or inflammation (white blood cell count [leukopenia (low) or normal], platelet count [thrombocytosis (high) or normal], C-reactive protein [high or normal], and erythrocyte sedimentation rate [high or normal]); and (4) vasculopathy (livedo reticularis [present or normal], periungual redness and swelling [present or normal], onychodermal band [abnormally prominent or normal], palatal petechiae [present or normal], von Willebrand factor antigen [high or normal], and D-dimer [high or normal]). We also report signs of anemia (hemoglobin [low or normal], mean corpuscular volume [low or normal], transferrin saturation [low or normal], and total iron binding capacity [high or normal]). Abnormalities were determined using each laboratory’s reference ranges. We report the first laboratory data collected during a period of neuropsychiatric symptom exacerbation and prior to the initiation of immunomodulatory therapy.

### 2.7 | Assessment of Pain and Sensory Disturbances

We assess pain and sensory characteristics at the time of arthritis diagnosis using the 31-Item PANS Scale [19] and the revised 2016 ACR fibromyalgia tool (Appendix A.6) [20].

### 2.8 | Statistical Analysis

We report descriptive statistics for demographics (Table 1), symptoms at clinical intake (Table A2), arthritis subtypes (Table 2), autoimmune and immunologic conditions (Table 2), musculoskeletal and nail characteristics (Table 3), ultrasonographic findings (Table 3), and signs of immune activation (Table 4). We estimated the cumulative incidence of autoimmune conditions including JIA and calculated 95% CIs using product-limit (Kaplan-Meier) survival analysis methods to account for censoring (Figure 1, Table A4). Analyses were conducted using lifelines and pandas in Python 3.10 [21,22]. Figure 1 was generated using matplotlib [23].

**FIGURE 1.**
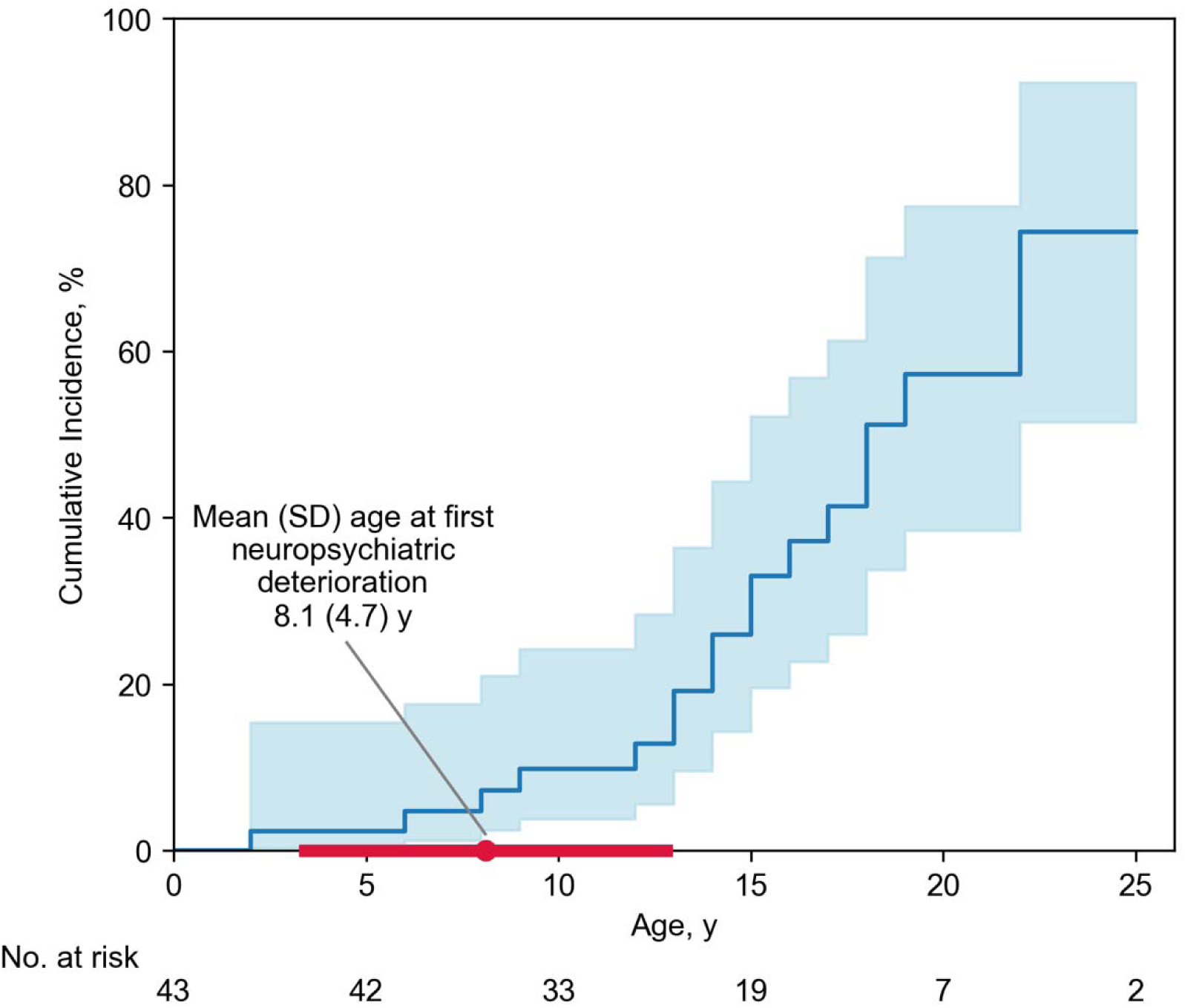
| Kaplan-Meier Plot, Cumulative Incidence of Autoimmune Conditions (Including Arthritis) Among Patients with ASD and Suspected Post-Infectious Neuropsychiatric Deteriorations. The shaded areas indicate the 95% CI. Abbreviations: ASD, autism spectrum disorder.

**TABLE 1.** | Demographics and Neuropsychiatric Presentation of Consecutive Patients with ASD and Suspected Post-Infectious Neuropsychiatric Deteriorations Seen at a Single Center.

| Characteristic | Patients (n=43) |
| --- | --- |
| Sex, No. (%) |  |
| Female | 13 (30) |
| Male | 30 (70) |
| Race, No. (%) |  |
| White or European | 33 (77) |
| Asian or Asian American | 4 (9) |
| Hispanic or Latino<sup>a</sup> | 1 (2) |
| Multiracial | 5 (12) |
| Age at first reported neuropsychiatric deterioration, mean (SD), years | 8.1 (4.7) |
| Age at deterioration prompting referral to clinic, mean (SD), years | 10.6 (4.4) |
| Age at ASD diagnosis, mean (SD), years | 8.6 (5.7) |
| Age at IBH Clinic presentation, mean (SD), years | 12.0 (4.0) |
| Age at first autoimmune condition (non-arthritis), mean (SD), years (n=9) | 12.7 (4.9) |
| Age at arthritis diagnosis, mean (SD), years (n=15) | 16.2 (5.5) |
| First neuropsychiatric deterioration preceded ASD diagnosis, No. (%) | 25 (58) |
| IBH Clinic follow-up time, mean (SD), years | 3.8 (3.7) |

*Abbreviations:* ASD, autism spectrum disorder; IBH, Immune Behavioral Health.
<sup>a</sup>May include White or European.

**TABLE 2.** | Arthritis Subtypes and Other Autoimmune or Immunologic Conditions Among Consecutive Patients with ASD and Suspected Post-Infectious Neuropsychiatric Deteriorations.

| **Characteristic** | **Patients (n=43)** |
| --- | --- |
| **Arthritis subtypes** |  |
| Any chronic arthritis | 15 (35) |
| Age at arthritis diagnosis, mean (SD), years (n=15) | 16.2 (5.5) |
| Subtype based on ILAR criteria, No. (%) |  |
| Enthesitis-related arthritis | 9 (21) |
| Psoriatic arthritis | 4 (9) |
| Oligoarticular arthritis |  |
| Persistent | 1 (2) |
| Extended | 0 |
| RF-positive polyarthritis | 0 |
| RF-negative polyarthritis | 0 |
| Undifferentiated arthritis | 1 (2) |
| Subtype based on ASAS criteria |  |
| Spondyloarthritis, No. (%) | 1 (2) |
| Peripheral, No./Total no. | 1/1 |
| Axial, No./Total no. | 0/1 |
| **Autoimmune and immunologic conditions (beyond arthritis)** |  |
| Any autoimmune condition, No. (%)<sup>a</sup> | 9 (21) |
| Age at diagnosis, mean (SD), years (n=9) | 12.7 (4.9) |
| Celiac disease | 3 (7) |
| Psoriasis | 2 (5) |
| Thyroiditis | 2 (5) |
| Inflammatory bowel disease<sup>b</sup> | 2 (5) |
| Systemic lupus erythematosus | 1 (2) |
| Any autoimmune condition above and arthritis | 6 (14) |
| Any other immunologic condition, No. (%)<sup>a</sup> | 25 (58) |
| Asthma | 12 (28) |
| Environmental or seasonal allergies | 14 (33) |
| Food allergies<sup>c</sup> | 9 (21) |
| Eosinophilic esophagitis | 3 (7) |
| Primary immunodeficiency | 3 (7) |
| Any other immunologic condition above and arthritis | 10 (23) |

*Abbreviations:* ASD, autism spectrum disorder; ILAR, International League of Associations for Rheumatology; RF, rheumatoid factor; ASAS, Assessment of SpondyloArthritis International Society.
<sup>a</sup>Criteria for classifying patients with these conditions are provided in Appendix A.5.
<sup>b</sup>Includes Crohn's disease and ulcerative colitis.
<sup>c</sup>This does not include "food sensitivities", which were present in 15 patients.

**TABLE 3.** | Musculoskeletal and Nail Characteristics Among 43 Consecutive Patients with ASD Referred for Post-Infectious Neuropsychiatric Deterioration, Stratified by Juvenile Idiopathic Arthritis (JIA) Criteria.

| Characteristic | Met JIA criteria | Did not meet JIA criteria |
| --- | --- | --- |
| **Musculoskeletal and nail characteristics (n=43)** |  |  |
| No. | 15 | 28 |
| **Inflammatory back pain (IBP)** |  |  |
| History of back pain, No./Total no. (%) | 6/15 (40) | 1/28 (4) |
| Meets Calin criteria for IBP, No./Total no. (%) | 5/15 (33) | 0 |
| **Physical examination (n=43)** |  |  |
| Prevalence of tenderness, No./Total no. (%)<sup>a</sup> |  |  |
| Distal interphalangeal joints | 9/15 (60) | 7/28 (25) |
| Sacroiliac joint tenderness | 9/15 (60) | 3/28 (11) |
| Spinous process tenderness (enthesitis) | 11/15 (73) | 4/28 (14) |
| Achilles tendon insertion (heel enthesitis) | 9/15 (60) | 4/28 (14) |
| Nail pitting, No./Total no. (%) | 9/15 (60) | 2/28 (7) |
| ≥1 one physical exam finding (above), No./Total no. (%) | 14/15 (93) | 11/28 (39) |
| **Joint ultrasonography (n=23)** |  |  |
| No. | 14 | 10 |
| Normal, No./Total no. (%) | 0 | 3/10 (30) |
| Abnormal, No./Total no. (%) | 14/14 (100) | 7/10 (70) |
| Abnormal findings on ultrasound, No./Total no. (%)<sup>a</sup> |  |  |
| Joint effusion | 14/14 (100) | 7/10 (70) |
| Synovial thickening or proliferation (synovitis) | 11/14 (79) | 2/10 (20) |
| Capsular thickening (capsulitis) | 5/14 (36) | 0 |
| Tenosynovitis<sup>b</sup> | 4/14 (29) | 0 |
| Ganglion cyst | 2/14 (14) | 0 |
| Bone erosion | 1/14 (7) | 0 |
| Nodules<sup>c</sup> | 0 | 1/10 (10) |
| Joints concerning for arthritis on ultrasound, No./Total no. (%)<sup>a</sup> |  |  |
| Wrist | 2/14 (14) | 0 |
| Fingers | 10/14 (71) | 2/10 (20) |
| Ankle | 4/14 (29) | 0 |
| Toes | 10/14 (71) | 5/10 (50) |

*Abbreviations:* ASD, autism spectrum disorder.
<sup>a</sup>Patients may fall into two or more categories.
<sup>b</sup>Tendon thickening with or without peritendinous or enthesal effusion
<sup>c</sup>One patient with subcutaneous nodule was also suspicious for ARF (see Appendix D.1).

**TABLE 4.** | Laboratory Markers and Physical Signs Which May Reflect Immune Activation Assessed at First Neuropsychiatric Symptom Exacerbation Captured in Clinic.

| Marker | No./Total no. tested (%) |
| --- | --- |
| **Autoimmune markers (nonspecific) (n=36)** |  |
| Anti-nuclear antibody, titer $\geq 1:160$ | 3/30 (10) |
| Anti-histone antibody, high | 4/8 (50) |
| Anti-thyroid antibody, high<sup>a</sup> | 6/18 (33) |
| Any positive result ($\geq 1$ marker) | 12/36 (33) |
| **Complement markers (nonspecific) (n=32)** |  |
| C1q binding assay, high | 5/25 (20) |
| Complement 3, low | 0/28 (0) |
| Complement 4, low | 10/31 (32) |
| Any positive result ($\geq 1$ marker) | 13/32 (41) |
| **Immune dysregulation or inflammation markers (nonspecific) (n=37)** |  |
| Leukopenia (WBC < 4000 cells/mL) | 2/37 (5) |
| Thrombocytosis (platelet count > 4000 x 10<sup>3</sup> cells/mL) | 1/37 (3) |
| C-reactive protein, high | 0/21 (0) |
| Erythrocyte sedimentation rate, high | 1/19 (5) |
| Any positive result ($\geq 1$ marker) | 4/37 (11) |
| **Vasculopathy (nonspecific) (n=43)** |  |
| von Willebrand factor antigen, high | 3/22 (14) |
| D-dimer, high | 0/16 (0) |
| Livedo reticularis, present | 7/43 (16) |
| Periungual erythema, present | 4/43 (9) |
| Onychodermal band, abnormally prominent | 12/43 (28) |
| Palatal petechiae, present | 1/43 (2) |
| Any positive result ($\geq 1$ marker) | 13/43 (30) |
| **At least one marker above** | 26/43 (60) |
| **At least two markers above** | 16/43 (37) |
| **Anemia markers (nonspecific) (n=33)** |  |
| Hemoglobin, low | 8/31 (26) |
| Mean corpuscular volume (MCV), low | 4/30 (13) |
| Iron binding, low<sup>b</sup> | 8/18 (44) |
| Any positive result ($\geq 1$ marker) | 14/33 (42) |

<sup>a</sup>Anti-thyroglobulin or anti-thyroid peroxidase antibodies.
<sup>b</sup>Low iron binding indicated by low transferrin saturation or high TIBC.

## 3 | RESULTS

The analyses included 43 patients with ASD (30 boys [70%]; 33 White/European [77%], 4 Asian/Asian American [9%], and 6 other or multiracial [14%]) (Table 1). The mean (SD) age at first clinic visit was 12.0 (3.9) years, with a mean (SD) follow-up time of 3.8 (3.7) years at time of analyses. The mean (SD) age at first neuropsychiatric deterioration was 8.1 (4.7) years, and the mean (SD) age at neuropsychiatric deterioration prompting clinic referral was 10.6 (4.4) years. Approximately half (58% [25 of 43]) carried the diagnosis of ASD before the deterioration that prompted clinic referral. The remainder (42% [18 of 43]) were referred for evaluation of ASD after this deterioration.

Fifteen patients met criteria for JIA with the following subtypes: Enthesitis-related arthritis (ERA) (60% [9 of 15]), psoriatic arthritis (PsA) (27% [4 of 15]), oligoarticular or undifferentiated (13% [2 of 15]) (Table 2). One patient with ERA also met ASAS criteria for peripheral spondylarthritis (SpA). The mean (SD) age at arthritis diagnosis was 16.2 (5.5) years.

Nine patients (21%) developed at least one non-JIA autoimmune condition: Celiac disease (7% [3 of 43]), psoriasis (5% [2 of 43]), thyroiditis (5% [2 of 43]), IBD (5% [2 of 43]), and SLE (2% [1 of 43]) (Table 2). Of these, six patients (14% of total cohort) also met criteria for JIA. All six were diagnosed with a non-JIA autoimmune condition before (12% [5 of 43]) or at the same time as (2% [1 of 43]) JIA. The cumulative incidence of autoimmune conditions (including JIA) by age 14 was 26% (95% CI, 14.3%-44.4%) (Figure 1, Table A4). We report pain and sensory characteristics at the time of arthritis diagnoses (Table A3).

Twenty-five patients (58%) had at least one immunologic condition: asthma (28% [12 of 43]), environmental or seasonal allergies (33% [14 of 43]), food allergies (21% [9 of 43]), EoE (7% [3 of 43]), and PID (7% [3 of 43]) (Table 2). Of these, 10 patients (40%) also developed JIA. Fifteen patients (35%) reported food sensitivities.

All IBP patients meeting Calin criteria [16] also met criteria for JIA and represented 33% (5 of 15) of the JIA cohort (Table 3). A limited exam of two joint categories (distal interphalangeal [DIP] and sacral iliac [SI] joints) and two entheseal sites (Achilles insertion and spinous processes [SP]) revealed tenderness in 93% of JIA patients (14 of 15) and 32% of non-JIA patients (9 of 28). Sixty percent of JIA patients (9 of 15) and 7% of non-JIA patients had nail pitting.

Twenty-four patients (56%) underwent joint ultrasonography, including 14 with JIA (Table 3). All JIA patients with joint ultrasonography had findings confirming arthritis, including effusions (100% [14 of 14]) and synovitis (79% [11 of 14]). Among 10 non-JIA patients with ultrasonography (36% [10 of 28]), seven (70%) had arthritis findings including effusions (70% [7 of 10]) and synovitis (20% [2 of 10]). The joints most frequently concerning for arthritis were fingers (71% [10 of 14] JIA; 20% [2 of 10] non-JIA) and toes (71% [10 of 14] JIA; 50% [5 of 10] non-JIA).

We report proportions of patients with markers of systemic inflammation in four subcategories (Table 4). Twenty-six patients (60%) had at least one marker (Table 4). Frequencies are calculated from the number of patients with data in each subcategory: autoimmunity (33% [12 of 36]), complement activation (41% [13 of 32]), immune dysregulation or inflammation (11% [4 of 37]), and vasculopathy (30% [13 of 43]). Twelve patients (28%) had markers in multiple subcategories. Fourteen patients had anemia (42%).

## 4 | DISCUSSION

Patients are referred to the IBH Clinic when a pediatrician suspects post-infectious neuropsychiatric deterioration. Clear deterioration(s) preceded the diagnosis of ASD in 58% of patients (25 of 43). The diagnosis of ASD was pursued in these patients given prior history of distinctive behaviors that increased with the deterioration prompting clinic referral.

### 4.1 | High Burden of Autoimmune Conditions in ASD

Consistent with prior studies [24–27], our cohort demonstrated elevated rates of autoimmune and immunologic conditions. However, these patients were evaluated in the context of suspected post-infectious neuropsychiatric deteriorations, which likely accounts for the higher rate of autoimmune conditions compared to other cohorts. JIA was the most frequent autoimmune condition, aligning with emerging literature suggesting that pain and rheumatologic illness may be under-recognized contributors to functional impairment in ASD [28–30]. Two patients were suspicious for acute rheumatic fever (ARF) but did not meet Jones criteria [31] and were therefore not classified as having a non-JIA autoimmune condition (Appendix D.1).

### 4.2 | Inflammatory Signs at First Neuropsychiatric Deterioration Observed in Clinic

While most patients did not have a classifiable autoimmune condition at clinic presentation, many patients displayed evidence of systemic inflammation during the first neuropsychiatric deterioration observed in clinic, with 37% (16 of 43) having multiple markers (Table 4). Eighteen patients (42%) had at least one marker of complement activation or autoimmunity. Thirty percent (13 of 43) had at least one marker of vasculopathy. These observations suggest that subclinical immune activation may be common in ASD during periods of neuropsychiatric decline.

### 4.3 | Anemia

Fourteen patients (33%) had at least one sign of anemia (Table 4), a known comorbidity of systemic inflammation. This frequency is comparable to other ASD cohorts [32–34]. Nutritional deficiency may not account for this finding; only one patient followed a vegan diet. However, food restriction (present in 51% [22 of 43]) may explain some of the anemia. Among patients without food restriction or a vegan diet (n=21), anemia was present in 5 of 15 (33%) who underwent testing.

### 4.4 | Arthritis Characteristics

ERA is a common childhood arthritis, comprising 3-11% of all pediatric arthritis [35], and is linked to infections. In PANS, ERA accounts for 67% of JIA cases [11,36]. ERA frequently evolves to PsA and SpA, and all three conditions are thought to be on a continuum with enthesitis being a key finding [37–39]. In this cohort, ERA and PsA were the most common subtypes, affecting 87% of JIA patients (13 of 15). One patient had persistent oligoarticular arthritis which may evolve to ERA, PsA, or SpA. One patient had undifferentiated arthritis (Appendix D.2). All JIA cases were thought to be on the spectrum of ERA.

DIP involvement is rare in most JIA forms (<10%) but more common in PsA [40–43]. Likewise, nail pitting – a rare finding – is more frequent in PsA, particularly among patients with DIP involvement [44,45]. Although only four patients (9%) had PsA, 21 (49%) had DIP tenderness and/or nail pitting. Twelve of 15 JIA patients (80%) had DIP involvement and/or nail pitting. The high rate of periungual redness may also reflect a related manifestation of PsA.

Patients with psoriasis and PsA often have psychiatric comorbidities [46,47]. In this cohort, the most common psychiatric symptoms at clinical presentation were anxiety (84% [36 of 43]), irritability (81% [35 of 43]), moodiness/depression (81% [35 of 43]), and obsessions/compulsions (77% [33 of 43]) (Table A2). Interestingly, among patients with obsessive compulsive disorder (OCD), there is an increased risk of psoriasis and other skin conditions [48]. Psoriasis and OCD may be linked through immunological mechanisms involving inflammation and dysregulated stress response [49], but this relationship remains debated [50].

### 4.5 | Arthritis Signs in Patients Not Meeting JIA Criteria

Among 28 patients not meeting JIA criteria (Appendix A.3), 10 (36%) had joint ultrasonography, with 7 (70%) showing signs of arthritis. Most failed to meet criteria because they either lacked persistent arthritis symptoms (pain and/or stiffness for >6 weeks) or because these symptoms were not articulated, possibly because psychiatric symptoms overshadowed joint complaints. Many of these patients received immunomodulatory therapy when synovitis or other inflammatory signs (labs, vascular signs) were present (manuscript in progress).

### 4.6 | Imaging for Arthritis Detection and Confirmation

Because pain and sensory dysregulation are common in ASD [51], we used imaging to confirm cases of JIA. In cases where the patient was known to under-report pain (i.e. minimal pain behavior in the setting of trauma), screened for arthritis using ultrasound. Because families were often overwhelmed by neuropsychiatric symptoms at initial visits, joint pain/stiffness were rarely reported. On specific questioning, however, families frequently describe stiff gait in morning, constant repositioning with prolonged sitting, and “gelling phenomena” after long car rides (difficulty straightening knees when getting out of car). While only one patient was diagnosed with SpA, 12% (5 of 43) met IBP criteria [16]. Among JIA patients, 93% (14 of 15) had imaging confirmation of joint inflammation, most commonly effusions (n=14) and synovitis (n=11). Only one patient had bone erosions, suggesting that we generally diagnosed arthritis in a pre-destructive stage.

### 4.7 | Comparison with PANS

In our prior PANS cohort, arthritis and autoimmunity were similarly prevalent [11,36]. The most common arthritis types (ERA, PsA, SpA) and features (DIP involvement, nail pitting) in PANS resemble those in this ASD cohort. However, our PANS cohort tends to be diagnosed with arthritis earlier, likely because PANS patients could better articulate joint pain/stiffness.

### 4.8 | Pain Dysregulation

Sensory dysregulation is common among children with ASD [51]. However, the extent and direction (hypo- or hyper-sensitivity) of these abnormalities vary [52,53]. Data from the 2016-2017 National Survey of Children’s Health show a high prevalence of pain among children with ASD [54].

To evaluate pain symptoms systematically, we prospectively administered the ACR fibromyalgia tool at each visit (Appendix A.6) [20]. Among 17 patients meeting threshold fibromyalgia scores, 14 (82%) had joint ultrasounds, all revealing inflammation. This suggests that the elevated pain scores were not occurring in isolation but were associated with objective inflammatory findings. Only three patients (7%) met full criteria for fibromyalgia, one of whom also met JIA criteria (ERA and SpA) (Table A3).

Average myofascial points around the time of arthritis diagnosis in our cohort was 6.2 (Table A3), compared with 4.8 reported in youth with fibromyalgia [55]. The reported level of pain was typically mild to moderate (73% [11 of 15]). Interestingly, fibromyalgia myofascial points overlap with enthesitis points [56]. Two patients with JIA reported no pain but the rheumatologist suspected arthritis due to warmth of joints.

Fibromyalgia and other pain disorders can complicate the assessment of arthritis. The features of fibromyalgia (widespread pain, fatigue, sleep and cognitive symptoms) overlap with SpA, and approximately 16% of patients with axial SpA also have fibromyalgia [57]. Patients with both conditions often receive higher doses of medications and frequently switch between biologics [58]. In this cohort, 53% of JIA patients (8 of 15) also carry a clinical diagnosis of pain amplification or fibromyalgia.

Given the difficulty of performing joint exams and evaluating pain among patients with ASD, ultrasound has been instrumental for evaluation of joint inflammation. We hypothesize that the current arthritis classification criteria – which requires that pain and joint tenderness be present – underrepresents cases of arthritis in ASD.

### 4.9 | Neurological Impacts of Systemic Inflammation

Systemic inflammation can lead to neuroinflammation and exacerbation of neuropsychiatric symptoms [59]. Our observations of elevated inflammatory markers and high prevalence of autoimmunity raise the possibility that systemic inflammatory processes could influence behavioral and cognitive changes.

## 5 | CONCLUSIONS

In this retrospective cohort study of youth with ASD who were referred due to suspected post-infectious neuropsychiatric deteriorations, we observed a high burden of inflammatory conditions including JIA, systemic autoimmune conditions, and other immunologic comorbidities. Many patients demonstrated objective signs of inflammation on laboratory testing and physical examination, even in the absence of formal autoimmune diagnoses. Musculoskeletal pain was frequently under-recognized due to overwhelming psychiatric burden at clinical presentation. These findings suggest that persistent immune dysregulation and inflammation may be common in youth with ASD who present with post-infectious neuropsychiatric deteriorations, and this dysregulation may contribute to joint inflammation, propensity to autoimmune conditions, and possibly neuropsychiatric symptoms since it has been recognized that systemic inflammation can contribute to neuropsychiatric changes [60]. Systematic evaluation for rheumatologic and inflammatory features could improve detection and treatment of such conditions. Future studies are warranted to clarify mechanisms linking immune activation, arthritis, and neuropsychiatric symptoms in ASD, and to determine whether targeted immunomodulatory interventions may improve outcomes.

## Supporting information

Supplementary Online Content (Appendix)

## Author Contributions

**M.M.:** conceptualization, methodology, data curation, writing—original draft preparation, writing—review and editing. **N.S.:** formal analysis, data curation, writing—original draft preparation, writing—review and editing, visualization. **J.S.:** conceptualization, methodology, data curation, writing—review and editing. **Z.S.:** data curation. **K.M.:** data curation. **C.M.:** data curation. **B.F.:** data curation, writing—review and editing. **K.A.:** data curation. **C.C.:** data curation. **A.A.:** data curation. **Y.X.:** data curation, writing—review and editing. **P.T.:** data curation, writing—review and editing. **M.S.:** data curation, writing—review and editing. **K.W.H.:** data curation, writing—review and editing. **M.T.:** data curation, writing—review and editing. **J.F.:** conceptualization, methodology, data curation, writing—original draft preparation, writing—review and editing, supervision, project administration, funding acquisition. All authors have read and agreed to the published version of the manuscript.

## Acknowledgments

We would like to thank the technicians in the Lucile Packard Children’s Hospital Stanford Department of Radiology for obtaining the images of the patients. We are especially grateful for the patients and families who participate in our research and our current and former members of our research staff, our Immune Behavioral Health Clinical Team, and our collaborating physicians.

## Institutional Review Board Statement

The Stanford Institutional Review Board (IRB) approved this retrospective chart review of all patients evaluated by the IBH Clinic (IRB#28533).

## Data Availability Statement

Deidentified participant data, study protocol, and statistical/analytic code will be shared on reasonable request from a qualified investigator after bidirectional data use agreements have been approved by the respective institutions.

## Abbreviations

ASD: autism spectrum disorder
SD: standard deviation
ERA: enthesitis-related arthritis
PsA: psoriatic arthritis
JIA: juvenile idiopathic arthritis
PANS: pediatric acute-onset neuropsychiatric disorder
IBP: inflammatory back pain
ACR: American College of Rheumatology
ILAR: International League of Associations for Rheumatology
ASAS: Assessment of SpondyloArthritis International Society
IBD: inflammatory bowel disease
SLE: systemic lupus erythematosus
EoE: eosinophilic esophagitis
PID: primary immunodeficiency
SpA: spondyloarthritis
DIP: distal interphalangeal
SI: sacral iliac
SP: spinous process
ARF: acute rheumatic fever
OCD: obsessive-compulsive disorder.

## REFERENCES

1. Bougeard, C.; Picarel-Blanchot, F.; Schmid, R.; Campbell, R.; Buitelaar, J. Prevalence of Autism Spectrum Disorder and Co-Morbidities in Children and Adolescents: A Systematic Literature Review. Front Psychiatry 2021, 12, 744709, doi:10.3389/fpsyt.2021.744709.

2. Underwood, J.F.G.; DelPozo-Banos, M.; Frizzati, A.; Rai, D.; John, A.; Hall, J. Neurological and Psychiatric Disorders among Autistic Adults: A Population Healthcare Record Study. Psychol Med 2023, 53, 5663–5673, doi:10.1017/S0033291722002884.

3. Mandell, D.S. Psychiatric Hospitalization among Children with Autism Spectrum Disorders. J Autism Dev Disord 2008, 38, 1059–1065, doi:10.1007/s10803-007-0481-2.

4. McMaughan, D.J.D.; Jones, J.L.; Mulcahy, A.; Tucker, E.C.; Beverly, J.G.; Perez-Patron, M. Hospitalizations Among Children and Youth With Autism in the United States: Frequency, Characteristics, and Costs. Intellect Dev Disabil 2022, 60, 484–503, doi:10.1352/1934-9556-60.6.484.

5. Mosner, M.G.; Kinard, J.L.; Shah, J.S.; McWeeny, S.; Greene, R.K.; Lowery, S.C.; Mazefsky, C.A.; Dichter, G.S. Rates of Co-Occurring Psychiatric Disorders in Autism Spectrum Disorder Using the Mini International Neuropsychiatric Interview. J Autism Dev Disord 2019, 49, 3819–3832, doi:10.1007/s10803-019-04090-1.

6. Balasco, L.; Provenzano, G.; Bozzi, Y. Sensory Abnormalities in Autism Spectrum Disorders: A Focus on the Tactile Domain, From Genetic Mouse Models to the Clinic. Front Psychiatry 2019, 10, 1016, doi:10.3389/fpsyt.2019.01016.

7. Mody, M.; Belliveau, J.W. Speech and Language Impairments in Autism: Insights from Behavior and Neuroimaging. N Am J Med Sci (Boston) 2013, 5, 157–161, doi:10.7156/v5i3p157.

8. Siniscalco, D.; Schultz, S.; Brigida, A.L.; Antonucci, N. Inflammation and Neuro-Immune Dysregulations in Autism Spectrum Disorders. Pharmaceuticals (Basel) 2018, 11, 56, doi:10.3390/ph11020056.

9. Chen, Y.; Du, X.; Zhang, X.; Li, F.; Yuan, S.; Wang, W.; Zhu, Z.; Wang, M.; Gu, C. Research Trends of Inflammation in Autism Spectrum Disorders: A Bibliometric Analysis. Front Immunol 2025, 16, 1534660, doi:10.3389/fimmu.2025.1534660.

10. Gagliano, A.; Cucinotta, F.; Giunta, I.; Di Modica, I.; De Domenico, C.; Costanza, C.; Germanò, E.; Frankovich, J. The Immune/Inflammatory Underpinnings of Neurodevelopmental Disorders and Pediatric Acute-Onset Neuropsychiatric Syndrome: A Scoping Review. Int J Mol Sci 2025, 26, 7767, doi:10.3390/ijms26167767.

11. Ma, M.; Masterson, E.E.; Gao, J.; Karpel, H.; Chan, A.; Pooni, R.; Sandberg, J.; Rubesova, E.; Farhadian, B.; Willet, T.;, et al. Development of Autoimmune Diseases Among Children With Pediatric Acute-Onset Neuropsychiatric Syndrome. JAMA Netw Open 2024, 7, e2421688, doi:10.1001/jamanetworkopen.2024.21688.

12. Harris, P.A.; Taylor, R.; Thielke, R.; Payne, J.; Gonzalez, N.; Conde, J.G. Research Electronic Data Capture (REDCap)--a Metadata-Driven Methodology and Workflow Process for Providing Translational Research Informatics Support. J Biomed Inform 2009, 42, 377–381, doi:10.1016/j.jbi.2008.08.010.

13. Harris, P.A.; Taylor, R.; Minor, B.L.; Elliott, V.; Fernandez, M.; O’Neal, L.; McLeod, L.; Delacqua, G.; Delacqua, F.; Kirby, J.;, et al. The REDCap Consortium: Building an International Community of Software Platform Partners. J Biomed Inform 2019, 95, 103208, doi:10.1016/j.jbi.2019.103208.

14. Office of Management and Budget Revisions to the Standards for Maintaining, Collecting, and Presenting Federal Data on Race and Ethnicity (OMB Directive No. 15).

15. Masterson, E.E.; Miles, K.; Schlenk, N.; Manko, C.; Ma, M.; Farhadian, B.; Chang, K.; Silverman, M.; Thienemann, M.; Frankovich, J.;, et al. Defining Clinical Course of Patients Evaluated for Pediatric Acute-Onset Neuropsychiatric Syndrome: Phenotypic Classification Based on 10 Years of Clinical Data. Dev Neurosci 2025, 47, 270–286, doi:10.1159/000545598.

16. Calin, A.; Porta, J.; Fries, J.F.; Schurman, D.J. Clinical History as a Screening Test for Ankylosing Spondylitis. JAMA 1977, 237, 2613–2614, doi:10.1001/jama.1977.03270510035017.

17. Petty, R.E.; Southwood, T.R.; Manners, P.; Baum, J.; Glass, D.N.; Goldenberg, J.; He, X.; Maldonado-Cocco, J.; Orozco-Alcala, J.; Prieur, A.-M.;, et al. International League of Associations for Rheumatology Classification of Juvenile Idiopathic Arthritis: Second Revision, Edmonton, 2001. J Rheumatol 2004, 31, 390–392.

18. Rudwaleit, M.; van der Heijde, D.; Landewé, R.; Akkoc, N.; Brandt, J.; Chou, C.T.; Dougados, M.; Huang, F.; Gu, J.; Kirazli, Y.;, et al. The Assessment of SpondyloArthritis International Society Classification Criteria for Peripheral Spondyloarthritis and for Spondyloarthritis in General. Ann Rheum Dis 2011, 70, 25–31, doi:10.1136/ard.2010.133645.

19. Bernstein, G.A.; Khan, M.H.; Freese, R.L.; Manko, C.; Silverman, M.; Ahmed, S.; Farhadian, B.; Ma, M.; Thienemann, M.; Murphy, T.K.;, et al. Psychometric Properties of the PANS 31-Item Symptom Rating Scale. J Child Adolesc Psychopharmacol 2024, 34, 157–162, doi:10.1089/cap.2023.0088.

20. Wolfe, F.; Clauw, D.J.; Fitzcharles, M.-A.; Goldenberg, D.L.; Häuser, W.; Katz, R.L.; Mease, P.J.; Russell, A.S.; Russell, I.J.; Walitt, B. 2016 Revisions to the 2010/2011 Fibromyalgia Diagnostic Criteria. Semin Arthritis Rheum 2016, 46, 319–329, doi:10.1016/j.semarthrit.2016.08.012.

21. Davidson-Pilon, C. Lifelines: Survival Analysis in Python. Journal of Open Source Software 2019, 4, 1317, doi:10.21105/joss.01317.

22. The pandas development team Pandas-Dev/Pandas: Pandas 2024.

23. Hunter, J.D. Matplotlib: A 2D Graphics Environment. Computing in Science & Engineering 2007, 9, 90–95, doi:10.1109/MCSE.2007.55.

24. Villarreal, V.R.; Katusic, M.Z.; Myers, S.M.; Weaver, A.L.; Nocton, J.J.; Voigt, R.G. Risk of Autoimmune Disease in Research-Identified Cases of Autism Spectrum Disorder: A Longitudinal, Population-Based Birth Cohort Study. J Dev Behav Pediatr 2024, 45, e46–e53, doi:10.1097/DBP.0000000000001232.

25. Doshi-Velez, F.; Avillach, P.; Palmer, N.; Bousvaros, A.; Ge, Y.; Fox, K.; Steinberg, G.; Spettell, C.; Juster, I.; Kohane, I. Prevalence of Inflammatory Bowel Disease Among Patients with Autism Spectrum Disorders. Inflamm Bowel Dis 2015, 21, 2281–2288, doi:10.1097/MIB.0000000000000502.

26. Lee, M.; Krishnamurthy, J.; Susi, A.; Sullivan, C.; Gorman, G.H.; Hisle-Gorman, E.; Erdie-Lalena, C.R.; Nylund, C.M. Association of Autism Spectrum Disorders and Inflammatory Bowel Disease. J Autism Dev Disord 2018, 48, 1523–1529, doi:10.1007/s10803-017-3409-5.

27. Lau, N.M.; Green, P.H.R.; Taylor, A.K.; Hellberg, D.; Ajamian, M.; Tan, C.Z.; Kosofsky, B.E.; Higgins, J.J.; Rajadhyaksha, A.M.; Alaedini, A. Markers of Celiac Disease and Gluten Sensitivity in Children with Autism. PLoS One 2013, 8, e66155, doi:10.1371/journal.pone.0066155.

28. Baeza-Velasco, C.; Cohen, D.; Hamonet, C.; Vlamynck, E.; Diaz, L.; Cravero, C.; Cappe, E.; Guinchat, V. Autism, Joint Hypermobility-Related Disorders and Pain. Front Psychiatry 2018, 9, 656, doi:10.3389/fpsyt.2018.00656.

29. Han, G.T.; Heavner, H.S.; Rains, T.R.; Hoang, A.H.; Stone, A.L. Chronic Pain in Autistic Youth: Clinical Prevalence and Reflections on Tailoring Evidence-Based Interventions from an Interdisciplinary Treatment Team. Children 2024, 11, 312, doi:10.3390/children11030312.

30. Donaghy, B.; Moore, D.; Green, J. Co-Occurring Physical Health Challenges in Neurodivergent Children and Young People: A Topical Review and Recommendation. Child Care in Practice 2023, 29, 3–21, doi:10.1080/13575279.2022.2149471.

31. Guidelines for the Diagnosis of Rheumatic Fever. Jones Criteria, 1992 Update. Special Writing Group of the Committee on Rheumatic Fever, Endocarditis, and Kawasaki Disease of the Council on Cardiovascular Disease in the Young of the American Heart Association. JAMA 1992, 268, 2069–2073.

32. Yang, W.; Liu, B.; Gao, R.; Snetselaar, L.G.; Strathearn, L.; Bao, W. Association of Anemia with Neurodevelopmental Disorders in a Nationally Representative Sample of US Children. The Journal of Pediatrics 2021, 228, 183–189.e2, doi:10.1016/j.jpeds.2020.09.039.

33. Latif, A.; Heinz, P.; Cook, R. Iron Deficiency in Autism and Asperger Syndrome. Autism 2002, 6, 103–114, doi:10.1177/1362361302006001008.

34. Al-Ali, S.F.; Russo, D.S.; Alkaissi, D.A. Association between Autism Spectrum Disorder and Iron Deficiency in Children Diagnosed Autism Spectrum Disorder in the Northern West Bank. 2015.

35. Ravelli, A.; Martini, A. Juvenile Idiopathic Arthritis. Lancet 2007, 369, 767–778, doi:10.1016/S0140-6736(07)60363-8.

36. Ma, M.; Sandberg, J.; Farhadian, B.; Silverman, M.; Xie, Y.; Thienemann, M.; Frankovich, J. Arthritis in Children with Psychiatric Deteriorations: A Case Series. Dev Neurosci 2023, 45, 325–334, doi:10.1159/000530854.

37. McGonagle, D.; Gibbon, W.; Emery, P. Classification of Inflammatory Arthritis by Enthesitis. Lancet 1998, 352, 1137–1140, doi:10.1016/S0140-6736(97)12004-9.

38. Polachek, A.; Li, S.; Chandran, V.; Gladman, D.D. Clinical Enthesitis in a Prospective Longitudinal Psoriatic Arthritis Cohort: Incidence, Prevalence, Characteristics, and Outcome. Arthritis Care Res (Hoboken) 2017, 69, 1685–1691, doi:10.1002/acr.23174.

39. McGonagle, D.; Aydin, S.Z.; Marzo-Ortega, H.; Eder, L.; Ciurtin, C. Hidden in Plain Sight: Is There a Crucial Role for Enthesitis Assessment in the Treatment and Monitoring of Axial Spondyloarthritis? Seminars in Arthritis and Rheumatism 2021, 51, 1147–1161, doi:10.1016/j.semarthrit.2021.07.011.

40. Hemke, R.; Nusman, C.M.; van der Heijde, D.M.F.M.; Doria, A.S.; Kuijpers, T.W.; Maas, M.; van Rossum, M.A.J. Frequency of Joint Involvement in Juvenile Idiopathic Arthritis during a 5-Year Follow-up of Newly Diagnosed Patients: Implications for MR Imaging as Outcome Measure. Rheumatol Int 2015, 35, 351–357, doi:10.1007/s00296-014-3108-x.

41. Sorokina, L.; Kaneva, M.; Artamonov, A.; Gordeeva, N.; Chikova, I.; Kostik, M. Clinical and Laboratory Features of Juvenile Idiopathic Arthritis with Wrist Involvement: Results of a Retrospective Cohort Study. World J Clin Pediatr 2024, 13, 91656, doi:10.5409/wjcp.v13.i3.91656.

42. Veale, D.; Rogers, S.; Fitzgerald, O. Classification of Clinical Subsets in Psoriatic Arthritis. Br J Rheumatol 1994, 33, 133–138, doi:10.1093/rheumatology/33.2.133.

43. Butbul, Y.A.; Tyrrell, P.N.; Schneider, R.; Dhillon, S.; Feldman, B.M.; Laxer, R.M.; Saurenmann, R.K.; Spiegel, L.; Cameron, B.; Tse, S.M.;, et al. Comparison of Patients with Juvenile Psoriatic Arthritis and Nonpsoriatic Juvenile Idiopathic Arthritis: How Different Are They? The Journal of Rheumatology 2009, 36, 2033–2041, doi:10.3899/jrheum.080674.

44. Butbul Aviel, Y.; Tyrrell, P.; Schneider, R.; Dhillon, S.; Feldman, B.M.; Laxer, R.; Saurenmann, R.K.; Spiegel, L.; Cameron, B.; Tse, S.M.;, et al. Juvenile Psoriatic Arthritis (JPsA): Juvenile Arthritis with Psoriasis? Pediatr Rheumatol 2013, 11, 11, doi:10.1186/1546-0096-11-11.

45. Lai, T.L.; Pang, H.T.; Cheuk, Y.Y.; Yip, M.L. Psoriatic Nail Involvement and Its Relationship with Distal Interphalangeal Joint Disease. Clin Rheumatol 2016, 35, 2031–2037, doi:10.1007/s10067-016-3319-5.

46. Hedemann, T.L.; Liu, X.; Kang, C.N.; Husain, M.I. Associations between Psoriasis and Mental Illness: An Update for Clinicians. General Hospital Psychiatry 2022, 75, 30–37, doi:10.1016/j.genhosppsych.2022.01.006.

47. Ferreira, B.I.R.C.; Abreu, J.L.P.D.C.; Reis, J.P.G.D.; Figueiredo, A.M.D.C. Psoriasis and Associated Psychiatric Disorders: A Systematic Review on Etiopathogenesis and Clinical Correlation. J Clin Aesthet Dermatol 2016, 9, 36–43.

48. Chou, Y.-J.; Tai, Y.-H.; Dai, Y.-X.; Lee, D.-D.; Chang, Y.-T.; Chen, T.-J.; Chen, M.-H. Obsessive–Compulsive Disorder and the Associated Risk of Autoimmune Skin Diseases: A Nationwide Population-Based Cohort Study. CNS Spectrums 2023, 28, 157–163, doi:10.1017/S1092852921000973.

49. Craver, A.E.; Chen, G.F.; Fan, R.; Levey, D.F.; Cohen, J.M. Association between Psoriasis and Obsessive-Compulsive Disorder: A Case-Control Study in the All of Us Research Program. Arch Dermatol Res 2024, 316, 280, doi:10.1007/s00403-024-03112-y.

50. Cosco, T.D.; Pillinger, T.; Emam, H.; Solmi, M.; Budhdeo, S.; Matthew Prina, A.; Maes, M.; Stein, D.J.; Stubbs, B.; Carvalho, A.F. Immune Aberrations in Obsessive-Compulsive Disorder: A Systematic Review and Meta-Analysis. Mol Neurobiol 2019, 56, 4751–4759, doi:10.1007/s12035-018-1409-x.

51. Baum, S.H.; Stevenson, R.A.; Wallace, M.T. Behavioral, Perceptual, and Neural Alterations in Sensory and Multisensory Function in Autism Spectrum Disorder. Prog Neurobiol 2015, 134, 140–160, doi:10.1016/j.pneurobio.2015.09.007.

52. Tordjman, S.; Anderson, G.M.; Botbol, M.; Brailly-Tabard, S.; Perez-Diaz, F.; Graignic, R.; Carlier, M.; Schmit, G.; Rolland, A.-C.; Bonnot, O.;, et al. Pain Reactivity and Plasma Beta-Endorphin in Children and Adolescents with Autistic Disorder. PLoS One 2009, 4, e5289, doi:10.1371/journal.pone.0005289.

53. Nader, R.; Oberlander, T.F.; Chambers, C.T.; Craig, K.D. Expression of Pain in Children with Autism. Clin J Pain 2004, 20, 88–97, doi:10.1097/00002508-200403000-00005.

54. Whitney, D.G.; Shapiro, D.N. National Prevalence of Pain Among Children and Adolescents With Autism Spectrum Disorders. JAMA Pediatr 2019, 173, 1203–1205, doi:10.1001/jamapediatrics.2019.3826.

55. Peterson, E.E.; Yao, C.; Sule, S.D.; Finkel, J.C. The Challenges of Identifying Fibromyalgia in Adolescents. Case Rep Pediatr 2022, 2022, 8717818, doi:10.1155/2022/8717818.

56. Roussou, E.; Ciurtin, C. Clinical Overlap between Fibromyalgia Tender Points and Enthesitis Sites in Patients with Spondyloarthritis Who Present with Inflammatory Back Pain. Clin Exp Rheumatol 2012, 30, 24–30.

57. Jones, G.T.; Mallawaarachchi, B.; Shim, J.; Lock, J.; Macfarlane, G.J. The Prevalence of Fibromyalgia in Axial Spondyloarthritis. Rheumatol Int 2020, 40, 1581–1591, doi:10.1007/s00296-020-04621-5.

58. Negm, A.; Alsaleh, J. THU0484 FIBROMYALGIA AND MULTIPLE SWITCHING OF BIOLOGICS IN SPONDYLOARTHRITIS. Annals of the Rheumatic Diseases 2020, 79, 479–480, doi:10.1136/annrheumdis-2020-eular.6224.

59. Sun, Y.; Koyama, Y.; Shimada, S. Inflammation From Peripheral Organs to the Brain: How Does Systemic Inflammation Cause Neuroinflammation? Front. Aging Neurosci. 2022, 14, doi:10.3389/fnagi.2022.903455.

60. Bauer, M.E.; Teixeira, A.L. Inflammation in Psychiatric Disorders: What Comes First? Annals of the New York Academy of Sciences 2019, 1437, 57–67, doi:10.1111/nyas.13712.

61. Aringer, M.; Costenbader, K.; Daikh, D.; Brinks, R.; Mosca, M.; Ramsey-Goldman, R.; Smolen, J.S.; Wofsy, D.; Boumpas, D.T.; Kamen, D.L.;, et al. 2019 European League Against Rheumatism/American College of Rheumatology Classification Criteria for Systemic Lupus Erythematosus. Arthritis & Rheumatology 2019, 71, 1400–1412, doi:10.1002/art.40930.

62. Weiss, P.F.; Brandon, T.G.; Aggarwal, A.; Burgos-Vargas, R.; Colbert, R.A.; Horneff, G.; Laxer, R.M.; Minden, K.; Ravelli, A.; Ruperto, N.;, et al. Classification Criteria for Axial Disease in Youth With Juvenile Spondyloarthritis. Arthritis Rheumatol 2024, 76, 1797–1808, doi:10.1002/art.42959.

