## Supplementary Online Content (Appendix) for "Autoimmunity and Arthritis in Youth with Autism and Suspected Post-Infectious Deteriorations"

**APPENDIX A | Additional Methodological Details**

**APPENDIX A.1** | Data Capture System (REDCap)

**APPENDIX A.2** | Date Assignments and Equivalencies for Determining Age at Events

**APPENDIX A.3** | Classification Criteria for Arthritis and Inflammatory Back Pain

**APPENDIX A.4** | Ultrasound Technology

**APPENDIX A.5** | Classification Criteria for Autoimmune Conditions, Other Immunologic Conditions, Food Sensitivities, and Pain Amplification

**APPENDIX A.6** | American College of Rheumatology (ACR) Fibromyalgia Tool

**APPENDIX B | Cohort Characteristics**

**TABLE A1** | Deterioration Characteristics of Consecutive Patients with ASD Referred for Post-Infectious Neuropsychiatric Deterioration, N=43

**TABLE A2** | Symptoms at Initial Clinical Presentation Among Consecutive Patients with ASD Referred for Post-Infectious Neuropsychiatric Deterioration, N=43

**TABLE A3** | Pain and Sensory Characteristics at the Time of Arthritis Diagnosis, N=15

**APPENDIX C | Cumulative Incidence of Autoimmune Conditions**

**TABLE A4** | Cumulative Incidence of Autoimmune Conditions Including Arthritis for Different Cumulative Time Points by Age (Years) Among Consecutive Patients with ASD Referred for Post-Infectious Neuropsychiatric Deterioration, N=43

**APPENDIX D | Case Descriptions**

**APPENDIX D.1** | Suspected Cases of Acute Rheumatic Fever (ARF)

**APPENDIX D.2** | Case of Undifferentiated Arthritis

This supplementary material has been provided by the authors to give readers additional information about their work.

**APPENDIX A | Additional Methodological Details**

**APPENDIX A.1** | Data Capture System (REDCap)

The Stanford REDCap platform is developed and operated by Stanford Medicine Research Technology team [12,13]. The REDCap platform services at Stanford are subsidized by a) Stanford School of Medicine Research Office, and b) the National Center for Research Resources and the National Center for Advancing Translational Sciences, National Institutes of Health, through grant UL1 TR003142.

**APPENDIX A.2** | Date Assignments and Equivalencies for Determining Age at Events

| **Phrase** | **Assigned (Estimated) Date** |
| --- | --- |
| In the spring | April 15 |
| In the summer | July 15 |
| In the fall / In autumn | October 15 |
| In the winter | January 15 |
| Middle of the week | The Wednesday of that week |
| Last week | The Wednesday of the week before |
| In the year / During the year | July 1 |
| Beginning of the year | January 1 |
| End of the year | December 31 |
| In the month / During the month | The 15th of that month |
| Beginning of the month | The 1st of that month |
| End of the month | The last day of that month |
| At age (years) | Birthday + age indicated + 6 months |
| At age (months) | Birthday + age indicated + 15 days |
| Within a range of dates | The median (middle) date within the range |

**APPENDIX A.3** | Classification Criteria for Arthritis and Inflammatory Back Pain

**Psoriatic Arthritis (PsA) (ILAR)** [17]

1. Arthritis and psoriasis, or
2. Arthritis and at least 2 of the following: Dactylitis, nail abnormalities (pitting or onycholysis), first-degree family history of psoriasis confirmed by a dermatologist

Exclusions: Presence of rheumatoid factor (RF) or systemic arthritis

**Enthesitis-Related Arthritis (ERA) (ILAR)** [17]

1. Arthritis and enthesitis, or
2. Arthritis or enthesitis with at least 2 of the following: Sacroiliac joint tenderness and/or inflammatory spinal pain, presence of HLA-B27, first- or second-degree family history of medically confirmed HLA-27-associated disease, anterior uveitis (usually associated with pain, redness, or photophobia), onset of arthritis in a boy after the age of 8 years

Exclusions: First- or second-degree family history of psoriasis confirmed by a dermatologist or presence of systemic arthritis as defined above

**Oligoarticular JIA (ILAR)** [17]

Arthritis affecting 1 to 4 joints during the first 6 months of disease.

Subcategories:

- Persistent oligoarthritis: Affects not more than 4 joints throughout disease course
- Extended oligoarthritis: Affects more than 4 joints after the first 6 months of disease

Exclusions: Psoriasis or first-degree family history of psoriasis, arthritis in an HLA-B27 positive male beginning after 6th birthday, ankylosing spondylitis, ERA, sacroiliitis with IBD, or acute anterior uveitis, or first-degree family history of one of these disorders, presence of IgM RF on at least 2 occasions at least 3 months apart, or presence of systemic JIA

**Rheumatoid Factor-Negative Polyarthritis (ILAR)** [17]

Arthritis affecting 5 or more joints during the first 6 months of disease and negative testing for RF

Exclusions: Psoriasis or first-degree family history of psoriasis, arthritis in an HLA-B27 positive male beginning after 6th birthday, ankylosing spondylitis, ERA, sacroiliitis with IBD, or acute anterior uveitis, or first-degree family history of one of these disorders, presence of IgM RF on at least 2 occasions at least 3 months apart, or presence of systemic JIA

**Rheumatoid Factor-Positive Polyarthritis (ILAR)** [17]

Arthritis affecting 5 or more joints during the first 6 months of disease and positive testing for RF (2 or more positive tests for RF at least 3 months apart during the first 6 months of disease)

Exclusions: Psoriasis or first-degree family history of psoriasis, arthritis in an HLA-B27 positive male beginning after 6th birthday, or ankylosing spondylitis, ERA, sacroiliitis with IBD, or acute anterior uveitis, or first-degree family history of one of these disorders

**Undifferentiated Arthritis (ILAR)** [17]

Arthritis that fulfills criteria in no category or in 2 or more of the above categories

**Axial Spondyloarthritis (ASAS)** [18]

1. Back pain for 3 or more months, and
2. Age at onset <45 years, and
3. Sacroiliitis on imaging (active acute inflammation on MRI highly suggestive of sacroiliitis associated with spondyloarthritis, or definite radiographic sacroiliitis), plus at least one of the following features: Inflammatory back pain, arthritis, heel enthesitis, uveitis, dactylitis, psoriasis, Crohn’s disease/ulcerative colitis, good response to NSAIDs, family history of spondyloarthritis, HLA-B27, elevated CRP

or

4. HLA-B27 plus at least 2 of the following: Inflammatory back pain, arthritis, heel enthesitis, uveitis, dactylitis, psoriasis, Crohn’s disease/ulcerative colitis, good response to NSAIDs, family history of spondyloarthritis, HLA-B27, elevated CRP

**Peripheral Spondyloarthritis (ASAS)** [18]

1. Arthritis or enthesitis or dactylitis, and
2. At least one of the following features: Psoriasis, IBD, preceding infection, HLA-B27, uveitis, sacroiliitis on imaging (radiographs or MRI)

or

3. At least 2 of the following: Arthritis, enthesitis, dactylitis, history of inflammatory back pain, family history of spondyloarthritis

**Inflammatory Back Pain (IBP) (Calin)** [16]

Patients must have 4 of the following:

1. Discomfort for 3 months or more
2. Back stiffness in morning
3. Age of onset is less than 40 years
4. Insidious onset of pain
5. Discomfort is relieved by exercise

*Abbreviations*: ILAR, International League of Associations for Rheumatology; HLA, human leukocyte antigen; IBD, inflammatory bowel disease; JIA, juvenile idiopathic arthritis; IgM, immunoglobulin M; ASAS, Assessment of SpondyloArthritis International Society; MRI, magnetic resonance imaging; NSAID, non-steroidal anti-inflammatory drug; CRP, c-reactive protein.

**APPENDIX A.4** | Ultrasound Technology

All musculoskeletal ultrasonographic procedures were conducted at the Stanford Radiology Division on either a GE LOGIQ E9 machine (GE Healthcare Inc., Princeton, NJ) utilizing a 6-15 MHz linear transducer or a Siemens ACYSON Sequoia machine (Siemens Healthineers AG, Erlangen, Germany) utilizing an 18 MHz linear transducer. Both greyscale and color Doppler images of the target joints were acquired and saved on the institute's Picture Archiving Communication System (PACS). For the fingers and toes, dedicated still images of the metacarpophalangeal and metatarsophalangeal joints were saved, while the proximal and distal interphalangeal joints were scanned and saved together. Results were interpreted by a board-certified radiologist (JS) with fellowship training at Boston Children’s Hospital (Harvard University) in musculoskeletal imaging.

**APPENDIX A.5** | Classification Criteria for Autoimmune Conditions, Other Immunologic Conditions, Food Sensitivities, and Pain Amplification

**Celiac Disease**

Must be diagnosed by a pediatric gastroenterologist and documented in the EMR. Must have either biopsy-proven disease or high-risk serotype (HLA-DQ2 or HLA-DQ8) coupled with high tissue transglutaminase antibodies.

**Psoriasis**

Must be diagnosed by a dermatologist and documented in the EMR.

**Thyroiditis**

Must be diagnosed by a pediatrician or pediatric endocrinologist (documented in the EMR) and have at least one of the following:

1. An ultrasound showing thyroid fullness
2. Elevated thyroid antibodies (anti-thyroid peroxidase [TPO] or anti-thyroglobulin) on two separate occasions

**Inflammatory Bowel Disease (IBD)**

Must be diagnosed by a pediatric gastroenterologist (documented in the EMR) and have endoscopic evidence of IBD.

**Systemic Lupus Erythematosus (SLE)** [61]

Must be classified as SLE by a pediatric rheumatologist using the EULAR/ACR classification criteria for SLE: Patient must have a positive ANA (≥ 1:80, entry criterion) with at least one clinical criterion and ≥ 10 points accumulated across multiple domains. Each clinical (constitutional, hematologic, neuropsychiatric, mucocutaneous, serosal, musculoskeletal, renal) and immunologic (antiphospholipid antibodies, complement proteins, SLE specific antibodies) domain contain a weighted number. The highest weighted criterion in each domain is counted toward the total score. Occurrence of a criterion on at least one occasion is sufficient. Criteria need not occur simultaneously.

**Asthma**

Must be documented in the EMR.

**Environmental or Seasonal Allergies**

Must be documented in the EMR.

**Food Allergies**

Must be diagnosed by an allergy or immunology specialist and documented in the EMR.

**Eosinophilic Esophagitis (EoE)**

Must be diagnosed by a pediatric allergist or gastroenterologist (documented in the EMR) using the following criteria:

1. Symptoms of esophageal dysfunction, or concomitant atopic conditions, or endoscopic findings of rings, furrows, exudates, edema, stricture, narrowing, and crepe-paper mucosa, and
2. Greater than or equal to 15 eos/hpf (~60 eos/mm2) on esophageal biopsy, and

Exclusion of non-EoE disorders that cause or potentially contribute to EoE.

**Primary Immunodeficiency (PID)**

Must be diagnosed by a pediatric immunologist and documented in the EMR.

- Common variable immunodeficiency (CVID): Patient has markedly reduced serum IgG levels in combination with low levels of IgA and/or IgM (<2 SD below the mean), poor or absent response to immunizations, and an absence of any other defined immunodeficiency state.
- Specific polysaccharide antibody deficiency (SPAD): Patient has a poor response to Pneumovax, i.e., <70% of 23 protective serotypes in children 6 years or older, or <50% in children younger than 6 years.
- Hypogammaglobulinemia: Patient has a reduction in any serum immunoglobulin levels (IgG, IgA, or IgM).
- Selective IgA deficiency: Patient older than 4 years has an isolated deficiency of IgA (i.e., in the setting of normal levels of IgG and IgM) and other causes of hypogammaglobulinemia have been excluded.

**Food Sensitivities**

Must be mentioned at least once in any clinical note.

**Pain Amplification or Fibromyalgia**

Must be diagnosed by a pediatric rheumatologist or pain medicine specialist and documented in the EMR.

*Abbreviations*: EMR, electronic medical record; HLA, human leukocyte antigen; EULAR, European Alliance of Associations for Rheumatology; ACR, American College of Rheumatology; ANA, anti-nuclear antibody; eos/hpf, eosinophils per high-power field; IgG, immunoglobulin G; IgA, immunoglobulin A; IgM, immunoglobulin M.

**APPENDIX A.6** | American College of Rheumatology (ACR) Fibromyalgia Tool

The 2016 ACR criteria for fibromyalgia [20] require that the subject meet all three of the following criteria: 1) Generalized pain, defined as pain in at least 4 of 5 regions, is present; 2) Symptoms have been present at a similar level for at least 3 months; and 3) Meet threshold scores for Widespread Pain Index (WPI) and the Malaise score – also called Symptom Severity scale (SSS): WPI ≥ 7 and SSS ≥ 5 OR WPI ≥ 4 and SSS ≥ 9.

**APPENDIX B | Cohort Characteristics**

**TABLE A1** | Deterioration Characteristics of Consecutive Patients with ASD Referred for Post-Infectious Neuropsychiatric Deterioration, N=43

| **Characteristic** | **Patients (n=43)** |
| --- | --- |
| First Reported Neuropsychiatric Deterioration |  |
| Age at deterioration, mean (SD), years | 8.1 (4.7) |
| Acuity of onset, No. (%) |  |
| Hyperacute (<72 hours) | 11 (26) |
| Acute (3-6 days) | 6 (14) |
| Subacute (1-8 weeks) | 8 (19) |
| Insidious (>8 weeks) | 7 (16) |
| Unclear | 2 (5) |
| Not documented | 9 (21) |
| Start date type, No. (%) |  |
| Start date clearly documented | 25 (58) |
| Start date estimated^a^ | 18 (42) |
| Neuropsychiatric Deterioration Prompting Referral to Clinic |  |
| Age at deterioration, mean (SD), years | 10.6 (4.4) |
| Acuity of onset, No. (%) |  |
| Hyperacute (<72 hours) | 11 (26) |
| Acute (3-6 days) | 6 (14) |
| Subacute (1-8 weeks) | 6 (14) |
| Insidious (>8 weeks) | 7 (16) |
| Unclear | 1 (2) |
| Not documented | 12 (28) |
| Start date type, No. (%) |  |
| Start date clearly documented | 27 (63) |
| Start date estimated^a^ | 16 (37) |

*Abbreviations*: ASD, autism spectrum disorder.

^a^See Appendix A.2 for methods for assigning estimated dates based on clinical notes.

**TABLE A2** | Symptoms at Initial Clinical Presentation Among Consecutive Patients with ASD Referred for Post-Infectious Neuropsychiatric Deterioration, N=43

| **Characteristic** | **Patients (n=43)** |
| --- | --- |
| Symptoms, No. (%) |  |
| Obsessions or compulsions | 33 (77) |
| Food/fluid refusal or avoidance^a^ | 23 (53) |
| Anxiety including phobias, panic attacks, and separation anxiety | 36 (84) |
| Moodiness, mood swings or depression | 35 (81) |
| Emotional lability | 19 (44) |
| Irritability | 35 (81) |
| Aggression, violence, or rage | 31 (72) |
| Oppositional or defiant behaviors | 24 (56) |
| Hyperactivity or impulsivity | 19 (44) |
| Trouble paying attention | 30 (70) |
| Behavioral regression | 26 (60) |
| Worsening of school performance | 27 (63) |
| Worsening of handwriting, copying, or art (dysgraphia) | 13 (30) |
| Cognitive symptoms (difficulty thinking, brain fog, memory problems) | 20 (47) |
| Pain (headaches, abdominal pain, body pain) | 23 (53) |
| Sleep disturbances | 26 (60) |
| Daytime wetting or bedwetting (enuresis) | 13 (30) |
| Increased urinary frequency | 13 (30) |
| Sensory amplification | 26 (60) |
| Hallucinations or delusions | 13 (30) |
| Tics (motor or vocal) | 22 (51) |
| No. symptoms at presentation, mean (SD) | 11.8 (4.0) |
| Meets PANS criteria, No. (%) | 17 (40) |

Abbreviations: ASD, autism spectrum disorder; PANS, pediatric acute-onset neuropsychiatric syndrome.

^a^One patient had fluid refusal/avoidance in the absence of food refusal/avoidance.

**TABLE A3** | Pain and Sensory Characteristics at the Time of Arthritis Diagnosis, N=15

| **Characteristic** | **Patients (n=15)** |
| --- | --- |
| Time to or from arthritis diagnosis, mean (SD), days | 11.8 (23.1) |
| Patient neuropsychiatric status, No. (%) |  |
| In a clear neuropsychiatric deterioration | 6 (40) |
| In a recovered state (relative to previous deterioration) | 4 (27) |
| Unclear state | 5 (33) |
| Numbness | 2 (13) |
| Shooting, burning, stabbing pain or pain that feels like an electric shock | 1 (7) |
| 31-Item PANS Scale [19], Pain (headaches, abdominal pain, body pain), No, (%) |  |
| 4 - Extreme | 0 |
| 3 - Severe | 1 (7) |
| 2 - Moderate | 10 (67) |
| 1 - Mild | 1 (7) |
| 0 - None | 2 (13) |
| No data | 1 (7) |
| **American College of Rheumatology (ACR) Fibromyalgia Tool**^a^ |  |
| **ACR Fibromyalgia Widespread Pain Index (WPI)** |  |
| Total score (out of 19), median (IQR) | 4 (1-6) |
| Total number of major areas (out of 5), median (IQR) | 3 (0-3) |
| **ACR Fibromyalgia Malaise (SSS), No. (%)** |  |
| Fatigue, moderate to severe | 9 (60) |
| Waking un-refreshed, moderate to severe | 7 (47) |
| Cognitive symptoms, moderate to severe | 8 (53) |
| Headaches, present | 9 (60) |
| Pain/cramps in abdomen, present | 6 (40) |
| Depression, present | 6 (40) |
| Total Score (out of 12), median (IQR) | 6 (3-8) |
| ACR Fibromyalgia Tool, threshold scores met, No. (%) |  |
| Before or at the time of arthritis diagnosis | 10 (67) |
| Ever | 11 (73) |
| ACR Fibromyalgia Tool, full fibromyalgia criteria satisfied, No. (%) |  |
| Before or at the time of arthritis diagnosis | 1 (7) |
| Ever | 1 (7) |
| Myofascial tender point exam (out of 18), median (IQR) (n=14)^b^ | 4 (0-11) |
| Clinician diagnosed pain amplification disorder and/or fibromyalgia at any time in the follow-up period, No. (%)^c^ | 8 (53) |

*Abbreviations*: PANS, pediatric acute-onset neuropsychiatric syndrome.

^a^The ACR fibromyalgia tool [20] combines the Widespread Pain Index (WPI) and Malaise score – also called Symptom Severity scale (SSS) – to assess fibromyalgia symptom burden. To meet the score threshold, subjects must meet either a) WPI ≥ 7 and SSS ≥ 5 or b) WPI ≥ 4 and SSS ≥ 9. See Appendix A.6 for full criteria.

^b^Myofascial tender point examinations were performed on 14 of 15 subjects meeting criteria for arthritis. Some examinations were completed on a different date than the other metrics listed above. The mean (SD) time between the nearest myofascial tender point examination and the diagnosis of arthritis is 117 (131) days.

^c^Diagnosis of pain amplification disorder or fibromyalgia preceded the diagnosis of arthritis in 6 patients.

**APPENDIX C | Cumulative Incidence of Autoimmune Conditions**

**TABLE A4** | Cumulative Incidence of Autoimmune Conditions Including Arthritis for Different Cumulative Time Points by Age (Years) Among Consecutive Patients with ASD Referred for Post-Infectious Neuropsychiatric Deterioration, N=43

| **At age (years)** | **Number at risk** | **Number censored** | **Number of events** | **Cumulative incidence % (95% CI)** |
| --- | --- | --- | --- | --- |
| **0** | 43 | 0 | 0 | 0 (0, 0) |
| **1** | 43 | 0 | 0 | 0 (0, 0) |
| **2** | 43 | 0 | 1 | 2.3 (0.3, 15.4) |
| **3** | 43 | 0 | 1 | 2.3 (0.3, 15.4) |
| **4** | 42 | 1 | 0 | 2.3 (0.3, 15.4) |
| **5** | 42 | 1 | 0 | 2.3 (0.3, 15.4) |
| **6** | 41 | 0 | 1 | 4.7 (1.2, 17.5) |
| **7** | 40 | 1 | 0 | 4.7 (1.2, 17.5) |
| **8** | 37 | 0 | 1 | 7.3 (2.4, 20.9) |
| **9** | 35 | 1 | 0 | 9.9 (3.8, 24.2) |
| **10** | 33 | 1 | 0 | 9.9 (3.8, 24.2) |
| **11** | 32 | 1 | 0 | 9.9 (3.8, 24.2) |
| **12** | 30 | 0 | 1 | 12.9 (5.5, 28.3) |
| **13** | 25 | 1 | 0 | 19.2 (9.6, 36.4) |
| **14** | 22 | 1 | 0 | 25.9 (14.3, 44.4) |
| **15** | 19 | 1 | 0 | 33.0 (19.6, 52.1) |
| **16** | 16 | 0 | 1 | 37.2 (22.7, 56.8) |
| **17** | 15 | 0 | 1 | 41.4 (26.0, 61.2) |
| **18** | 10 | 1 | 0 | 51.1 (33.7, 71.3) |
| **19** | 8 | 0 | 1 | 57.2 (38.5, 77.4) |
| **20** | 7 | 1 | 0 | 57.2 (38.5, 77.4) |
| **21** | 6 | 1 | 0 | 57.2 (38.5, 77.4) |
| **22** | 4 | 0 | 1 | 74.3 (51.5, 92.3) |
| **23** | 3 | 1 | 0 | 74.3 (51.5, 92.3) |
| **24** | 2 | 1 | 0 | 74.3 (51.5, 92.3) |

*Abbreviations*: ASD, autism spectrum disorder

**APPENDIX D | Case Descriptions**

**APPENDIX D.1** | Suspected Cases of Acute Rheumatic Fever (ARF)

Two patients in our cohort were suspicious for acute rheumatic fever (ARF) but did not meet the Jones criteria [31]. One patient had a post-S. pyogenes neuropsychiatric deterioration and subcutaneous nodules that resolved over 8-12 months with treatment (amoxicillin and ibuprofen). The other patient suspicious for ARF had arthritis, but they did not fully meet the Jones criteria as they presented later in the course with resolving inflammation.

**APPENDIX D.2** | Case of Undifferentiated Arthritis

One patient in our cohort had undifferentiated arthritis. This patient had ultrasound findings of joint effusion, synovitis, and capsulitis, with DIP joint involvement. Capsulitis and DIP joint tenderness are seen in adults with PsA [42]. However, the patient did not have evidence of psoriasis, and we were able to assess for enthesitis. The patient also had a sclerotic focus on the ilium adjacent to the sacroiliac joint. This patient does not meet the new JIA SpA criteria [62], as we were unable to assess for pain.
